# Associations between alcohol use and accelerated biological ageing

**DOI:** 10.1101/2020.11.24.20237156

**Authors:** Sunniva M. K. Bøstrand, Kadi Vaher, Laura De Nooij, Mathew A. Harris, James H. Cole, Simon R. Cox, Riccardo E. Marioni, Daniel L. McCartney, Rosie M. Walker, Andrew M. McIntosh, Kathryn L. Evans, Heather C. Whalley, Robyn E. Wootton, Toni-Kim Clarke

## Abstract

**Background:** Harmful alcohol use is a leading cause of premature death, and is associated with age-related disease. Ageing is highly variable between individuals, and may deviate from chronological ageing, suggesting that biomarkers of biological ageing (based on DNA methylation or brain structural measures) may be clinically relevant. Here, we investigated the relationships between alcohol phenotypes and both brain and DNA methylation age estimates.

**Methods:** First, using data from UK Biobank and Generation Scotland, we tested the association between alcohol consumption (units/week) or hazardous use (AUDIT scores), and accelerated brain and epigenetic ageing in 20,258 and 8,051 individuals, respectively. Second, we used Mendelian randomization to test for a causal effect of alcohol consumption levels and alcohol use disorder (AUD) on biological ageing.

**Results:** Alcohol use showed a consistent positive association with higher predicted brain age (AUDIT-C: β=0.053, p=3.16×10^−13^; AUDIT-P: β=0.052, p=1.6×10^−13^; total AUDIT score: β=0.062, p=5.52×10^−16^; units/week: β=0.078, p=2.20×10^−16^), and DNA methylation GrimAge (Units/week: β=0.053, p=1.48×10^− 7^) and PhenoAge (Units/week: β=0.077, p=2.18×10^−10^). Mendelian randomization analyses revealed some evidence for a causal effect of AUD on accelerated brain ageing (β=0.272, p=0.044), and no evidence for a causal effect of alcohol consumption levels on accelerated biological ageing.

**Conclusions:** We provide consistent phenotypic evidence linking alcohol use with accelerated biological ageing. There is possible evidence for a causal effect of AUD on brain age, but not for any other alcohol-related trait on brain or epigenetic age acceleration. Future studies investigating the mechanisms associating alcohol use with accelerated biological ageing are warranted.

## Introduction

Harmful alcohol use is a leading cause for premature death globally (1). Excessive alcohol use affects multiple tissues (2,3), and is associated with an increased risk for all-cause mortality (4) and age-related diseases including diabetes, liver diseases, and dementia (5–7). While some studies have demonstrated that moderate alcohol consumption is negatively associated with mortality (8), a recent large scale epidemiological investigation has challenged this view, showing that even small amounts of alcohol negatively impact on health (1). Ageing itself is a complex process of progressive deterioration due to the accrual of cellular damage over time (9), and rates of these age-associated biological processes vary between individuals. This could account for some of the variation in susceptibility to age-related disease and suggests that measures of biological ageing may be more clinically relevant than chronological age (10,11). Several biomarkers of biological ageing have been proposed to measure individual variation in biological ageing, including those based on DNA methylation (DNAm) or brain magnetic resonance imaging (MRI). To date, studies investigating associations between alcohol use and biological ageing have been limited to small sample sizes, report inconsistent findings, and have not probed the potential causality of these associations.

Recent analytical approaches, such as that developed by Cole et al. (10), use machine learning algorithms trained to predict chronological age from brain structural MRI data. Testing these algorithms on new structural data produces a metric of brain age that correlates strongly with chronological age, while its deviation from chronological age reflects accelerated/decelerated brain ageing (12). Accelerated brain ageing predicts mortality in older adults and correlates with cognitive and physical decline (13). Several epigenetic clocks have also been used to characterise biological ageing and are hypothesised to capture molecular processes involved in declining tissue function (14). These biomarkers use weighted averages of methylation levels at specific cytosine-phosphate-guanine (CpG) sites to produce estimates of epigenetic age. Similarly to brain age, a greater positive deviation in DNAm age from chronological age predicts all-cause mortality (15,16), and has been linked to a range of age- and lifestyle-related health outcomes, including exercise and diet (17), BMI and obesity (18), cognitive ability (19,20), and Alzheimer’s disease (21).

Previous work has associated alcohol use with the acceleration of both brain and DNAm ageing (17,22–27). Ning et al (22) showed that more frequent consumption of alcohol was associated with a higher brain age relative to peers, with the lowest brain age found in individuals who reported drinking only occasionally. Higher alcohol intake frequency was also associated with an older-appearing brain using multiple MRI modalities to predict brain-age (27). Similarly, higher levels of alcohol consumption across 30 years of follow-up were associated with reductions in grey matter density and white matter structural integrity, suggesting that heavy alcohol consumption may lead to accelerated brain ageing (2).

Alcohol use is associated with variation in DNAm (28,29), thus, investigations into the associations between alcohol use and epigenetic ageing could allow insight into the shared molecular mechanisms underlying harmful alcohol use and ageing. Previous studies have reported a greater rate of epigenetic ageing in individuals with a diagnosis of alcohol dependence using two measures of epigenetic ageing; the Horvath clock, derived from DNAm levels at 353 CpG sites (14,23) and PhenoAge (24), a novel epigenetic biomarker of phenotypic age derived as a composite of chronological age and a selection of clinical characteristics representing death and disease risk (30). However, the associations between alcohol consumption levels and epigenetic ageing are complex, with studies showing positive (25), negative (17) and non-linear (26) relationships. Additionally, a recent study suggests both positive and negative genetic associations between various alcohol-related traits and epigenetic ageing using LD score regression (31).

Cross-sectional, observational epidemiological studies are prone to bias from residual confounding and reverse causation, limiting the investigations of cause-effect relationships. Genome-wide association studies (GWAS) have identified several loci implicated in both the clinical diagnosis of alcohol use disorder (AUD) and varying alcohol consumption levels (32,33). These genetic variants can be utilised as instruments to determine the most likely direction of effect between a modifiable exposure, alcohol use, and the outcome biological ageing using Mendelian Randomization (MR) (34,35). It follows the logic that if a modifiable exposure (e.g. alcohol use) is the cause of an outcome (e.g. accelerated biological ageing), then individuals with genetic variants predisposing them towards increased alcohol use should be more likely to experience accelerated biological ageing.

Here, we investigated the relationship between alcohol use and biological ageing, using the largest brain imaging (N=20,258) and DNAm (N=8,051) datasets to date. We hypothesised that higher levels of alcohol consumption would associate with both higher DNAm age and brain age, and that these associations would reflect a causal effect of higher alcohol use on accelerated biological ageing.

## Methods and Materials

### Study Populations: UK Biobank

UK Biobank (UKB) comprises N=502,617 individuals recruited from across the UK (36). UKB received ethical approval from the NHS National Research Ethics Service North West (reference:11/NW/0382). The present study was carried out under UKB project ID 4844. At the time of writing, we used the latest available UKB neuroimaging (see protocol in (36)) release consisting of N=21,386 individuals. After removing extreme outliers (defined as MRI measurements >5*SD from the mean), cases of image acquisition problems and excluding previous or never drinkers, N=20,258 individuals were included in the present study (see Table S1 and Figure S1 for demographics and sample selection).

#### Alcohol consumption

Lifestyle measures were collected at baseline and online follow-up. Alcohol use in units/week at baseline was calculated by converting the sum of reported average weekly intake of red wine, champagne plus white wine, beer plus cider, spirits, fortified wine and other alcoholic drinks into alcohol units. At online follow-up, a subset of participants (N=14,710) completed the Alcohol Use Disorders Identification Test (AUDIT), a 10-item screening tool developed by the World Health Organization (37) to assess alcohol consumption and alcohol-related behaviours and problems. To examine the relationship between alcohol use and brain age, we used four measures of alcohol consumption: Alcohol units/week and three measures from the AUDIT questionnaire - a composite score of alcohol consumption (AUDIT-C; sum of questions 1-3), problematic alcohol use (AUDIT-P; sum of questions 4-10) and the total score across all items (AUDIT-T).

#### Brain Age estimates

We utilized a measure of brain age derived from structural T1-weighted MRI data as described by Cole et al (10), and implemented using the brainageR software package (https://github.com/james-cole/brainageR) (38). In the sample of current drinkers, the correlation between brain age and chronological age was r=0.734, p<2.2×10^−16^ (full demographics in Table S1). This measure was subsequently residualised over chronological age, sex, imaging site (Manchester/Newcastle) and scanner head position X, Y and Z coordinates. The residualised brain age measure reflected deviation of brain age from chronological age (controlling for aforementioned covariates), with positive values representing accelerated brain ageing.

### Study populations: Generation Scotland

#### The Generation Scotland

Scottish Family Health Study (GS:SFHS) cohort comprises 23,690 individuals aged ≥18 years at recruitment and is described in detail elsewhere (39,40). At baseline, participants were assessed for a range of health, demographic and lifestyle factors, and provided samples for DNA extraction. GS:SFHS has been granted ethical approval from the NHS Tayside Committee on Medical Research Ethics, on behalf of the National Health Service (reference: 05/S1401/89) and has Research Tissue Bank Status (reference: 15/ES/0040). The present study includes individuals for whom information about alcohol consumption, smoking, BMI and DNAm data (profiled in two sets, see below) was acquired at baseline (full demographics in Table S2).

#### Alcohol consumption

We used self-reported units/week as a quantification of a person’s alcohol consumption. The present study includes individuals who reported being current drinkers; participants with alcohol consumption in units/week >4*SD from the mean were excluded from the analysis.

#### DNA methylation profiling

Whole blood DNAm was profiled using the Infinium MethylationEPIC BeadChip (Illumina Inc., San Diego, California) in 9778 participants across two processing sets (set 1 N=5,190 (comprised related individuals), set 2 N=4,583 (comprised unrelated (to each other and to set 1) individuals)) and quality controls were carried out using standard methods outlined in supplementary methods as described previously (41,42). The final DNAm dataset comprised data for 5,087 individuals in set 1 and 4,450 individuals in set 2. Participant data in the two sets were analysed separately and then meta-analysed. After applying exclusion criteria (see above) the current study included 4,260 participants from set 1 and 3,791 participants from set 2 (full demographics in Table S2).

#### Epigenetic estimates of age

Four DNAm-based estimates of age were calculated using the online age calculator (https://dnamage.genetics.ucla.edu/) developed by Horvath (14): Hannum (43) and Horvath (14) epigenetic age, DNAm GrimAge (44) and DNAm PhenoAge (30), which all strongly correlated with chronological age (Table S3). From these estimates, four age-adjusted epigenetic age acceleration (EAA) measures (intrinsic and extrinsic EAA (IEAA and EEAA, respectively) (45–47), AgeAccelGrim (44) and AgeAccelPheno (30)) were calculated (see Supplementary Methods). The age acceleration measures were uncorrelated with chronological age (Table S3).

### Statistical Methods

All statistical analyses were performed in R (versions 3.3.2, 3.6.1 and 4.0.1) (48). Scaling by z-transformation was applied for all numeric variables in the regression models.

#### Association between alcohol use and brain age in UK Biobank

The variables AUDIT-C, AUDIT-P, AUDIT total scores and alcohol consumption in units/week were entered separately into linear models to test for association with residualised brain age. Smoking status (coded at baseline as a binary value denoting whether individuals had ever smoked or never smoked), age and sex were added as covariates in each model in order to control for the effects of these variables. Results were plotted using the packages ggplot2 (49) and ggstance (50). Benjamini-Hochberg correction for false discovery rate (FDR) was applied to the regression models.

#### Association between alcohol consumption and epigenetic age acceleration in GS:SFHS

Set 1 data (related subset of GS:SFHS) statistical analyses were conducted in ASReml-R version 3.0 (www.vsni.co.uk/software/asreml) to fit a linear mixed model to control for relatedness within the sample by fitting an inverse relationship matrix derived from pedigree information as a random effect. Set 2 data (unrelated subset of GS:SFHS) was analysed using linear regression (lm) function in base R. In each model, EAA was fit as the dependent variable and alcohol consumption (as 1+log_10_(units/week) to adjust for non-normal distribution) as the independent variable; sex, BMI, smoking pack years (at baseline, individuals were asked to self-report their tobacco exposure (cigarettes/day), age when they started smoking, years since stopped smoking, and pack years variable was calculated as packs (20 cigarettes/pack) smoked per day multiplied by years as a smoker), and inverse relationship matrix fitted as a random effect only for set 1 in ASReml were added as covariates. As smoking is strongly associated with DNAm, sensitivity analyses were conducted in non-smoking individuals (i.e., who reported never having smoked tobacco); n=2,207 in set 1 and n=1,998 in set 2. To combine the coefficient estimates from the two sets into a single estimate, we applied an inverse variance-weighted fixed-effects meta-analysis model using the function ‘metagen’, implemented in the *meta* package in R (51). FDR correction for multiple testing was applied across all fully adjusted models (fully adjusted regressions in set 1 and 2, and meta-analysis) and separately for smoking sensitivity analysis. Results were plotted using the function ‘forest’ in *meta* package in R.

### Two-sample Mendelian randomization

Two-sample MR analysis was performed using the *TwoSampleMR* package from MRBase (52) using summary statistics extracted from non-overlapping GWASs (AUDIT-C and AUD GWAS: (33), AgeAccelGrim and AgeAccelPheno GWAS: (31), brain age GWAS: unpublished (see Supplementary methods)). See Supplementary Methods for the information about the exposure and outcome GWASs, and Tables S4 and S5 for the MR input testing causal effects of AUDIT-C and AUD, respectively, on biological ageing.

We performed two MR analyses with brain age as the outcome, where AUD or AUDIT-C were the exposures, respectively. One MR analysis was performed with either AgeAccelGrim or AgeAccelPheno as the outcome and AUDIT-C as the exposure. The main MR models included 13 SNPs to probe for the causal effect of AUDIT-C on biological ageing and 10 SNPs to probe for the causal effect of AUD on brain ageing.

We applied complementary methods of two-sample MR methods (inverse variance weighted (IVW), MR-Egger, weighted median, and weighted mode-based estimation). IVW was the main analysis with each of the others providing sensitivity analyses which each make different assumptions about the nature of pleiotropy (where the genetic variant associates with the outcome via an independent pathway to the exposure). Therefore, the strongest evidence for a causal effect would be where the estimates from all methods are consistent. To test the suitability of the MR-Egger method, the I^2^ statistic was calculated to quantify the degree of regression dilution bias due to measurement error of SNP-exposure effects (53). Where the I^2^ estimate (Table S6) was between 0.6 and 0.9, simulation extrapolation (SIMEX) correction (54) was applied to the MR-Egger analysis. The mean F-statistic as an indicator of instrument strength was also calculated (Table S7). Further, we used the MR-Egger intercept to test for the presence of horizontal pleiotropy (Table S9), Steiger filtering (55) to test for the most likely direction of effect (Tables S10-S13), and calculated Cochran’s Q (56) to assess heterogeneity suggestive of pleiotropy (Table S8). When there was evidence for a causal effect based on the IVW model, we performed MR Presso (57) and Radial MR (58) to detect potential outliers.

## Results

### Alcohol use associations with brain age in UK Biobank

Using linear regression we found a consistent positive relationship between brain age and the four measures of alcohol use (alcohol units/week (N=20,258, 52.1% female), AUDIT-C, AUDIT-P and AUDIT-T (N=14,710, 53.5% female)) (Figure 1), with higher levels of self-reported alcohol consumption associated with a more advanced brain age. The largest effect was found for alcohol consumption measured in units/week (β=0.078, 95% CI [0.066; 0.093], p<2.20×10^−16^).

**Figure 1.**
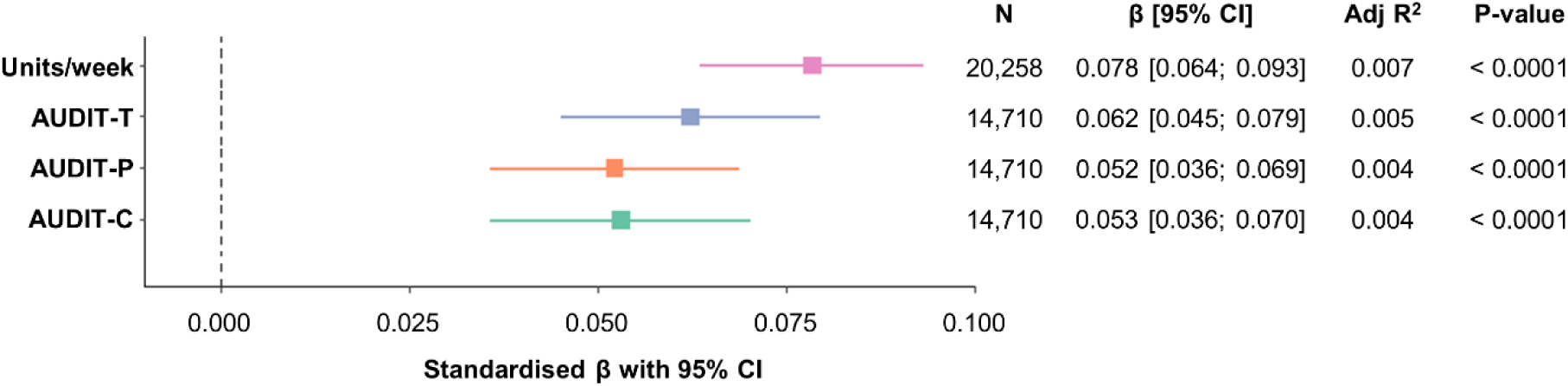
Alcohol use is associated with advanced brain age. Linear regression models predicting residual brain age from AUDIT-C, AUDIT-P, AUDIT-T and alcohol units, in current drinkers adjusted for smoking status. Plot shows standardised **β** coefficients with 95% confidence intervals. CI = confidence interval.

### Alcohol use associations with epigenetic age in GS:SFHS

Linear regression and fixed effect meta-analyses were used to investigate associations between alcohol consumption (units/week) and four measures of EAA (IEAA, EEAA, AgeAccelGrim and AgeAccelPheno) in 8,051 individuals in total (set 1 n=4,260 (60.9% female), set 2 n=3,791 (55.3% female)) from the GS:SFHS cohort (full demographics in Table S2). We found a positive association between alcohol consumption and AgeAccelGrim (β=0.053 [0.034; 0.071], p=1.48×10^−7^) and AgeAccelPheno (β=0.077 [0.055; 0.100], p=2.18×10^−10^), but not between alcohol consumption and IEAA or EEAA (Figure 2).

**Figure 2.**
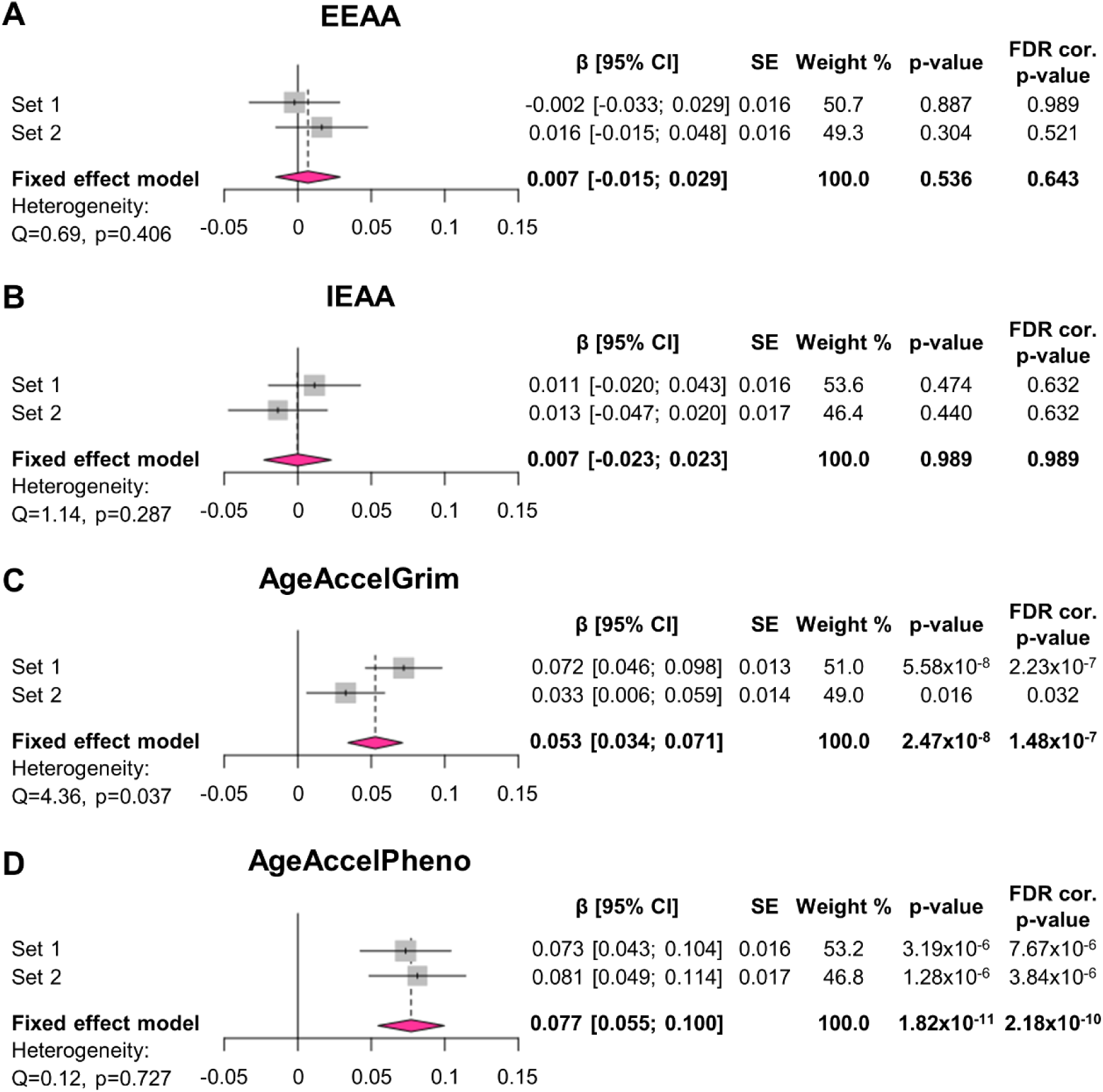
Alcohol consumption is associated with two measures of advanced epigenetic age. Effects of alcohol consumption (units/week) on (A) EEAA, (B) IEAA, (C) AgeAccelGrim and (D) AgeAccelPheno in fully adjusted models. Values on forest plot indicate standardised **β** with 95% confidence intervals. Models are adjusted for sex, BMI and pack years in sets 1 and 2, and relatedness in set 1 by fitting pedigree information as a random effect in general linear mixed models using advanced restricted maximum likelihood (ASReml) method. Fixed effect inverse variance weighted meta-analysis was applied using R package *meta* to combine the standardised coefficient estimates in set 1 and set 2. FDR correction was applied across all models in sets 1 and 2, and all meta-analysis models (12 models in total). Sample size: n=4260 in set 1, n=3791 in set 2 (n=8051 included in meta-analyses). EEAA = extrinsic epigenetic age acceleration, IEAA = intrinsic epigenetic age acceleration, SE = standard error, CI = confidence interval, FDR = false-discovery rate.

As smoking is strongly associated with DNAm, we conducted a sensitivity analysis by exploring the associations between alcohol consumption and AgeAccelGrim and AgeAccelPheno in a subset of non-smoking participants (Figure 3). The positive associations between alcohol consumption and the two EAA measures remained significant in non-smokers, but the effect size was slightly (∼15%) attenuated for AgeAccelGrim (β=0.045 [0.026; 0.061], p=6.48×10^−6^) while it remained similar for AgeAccelPheno (β=0.074 [0.043; 0.104], p=7.74×10^−6^).

**Figure 3.**
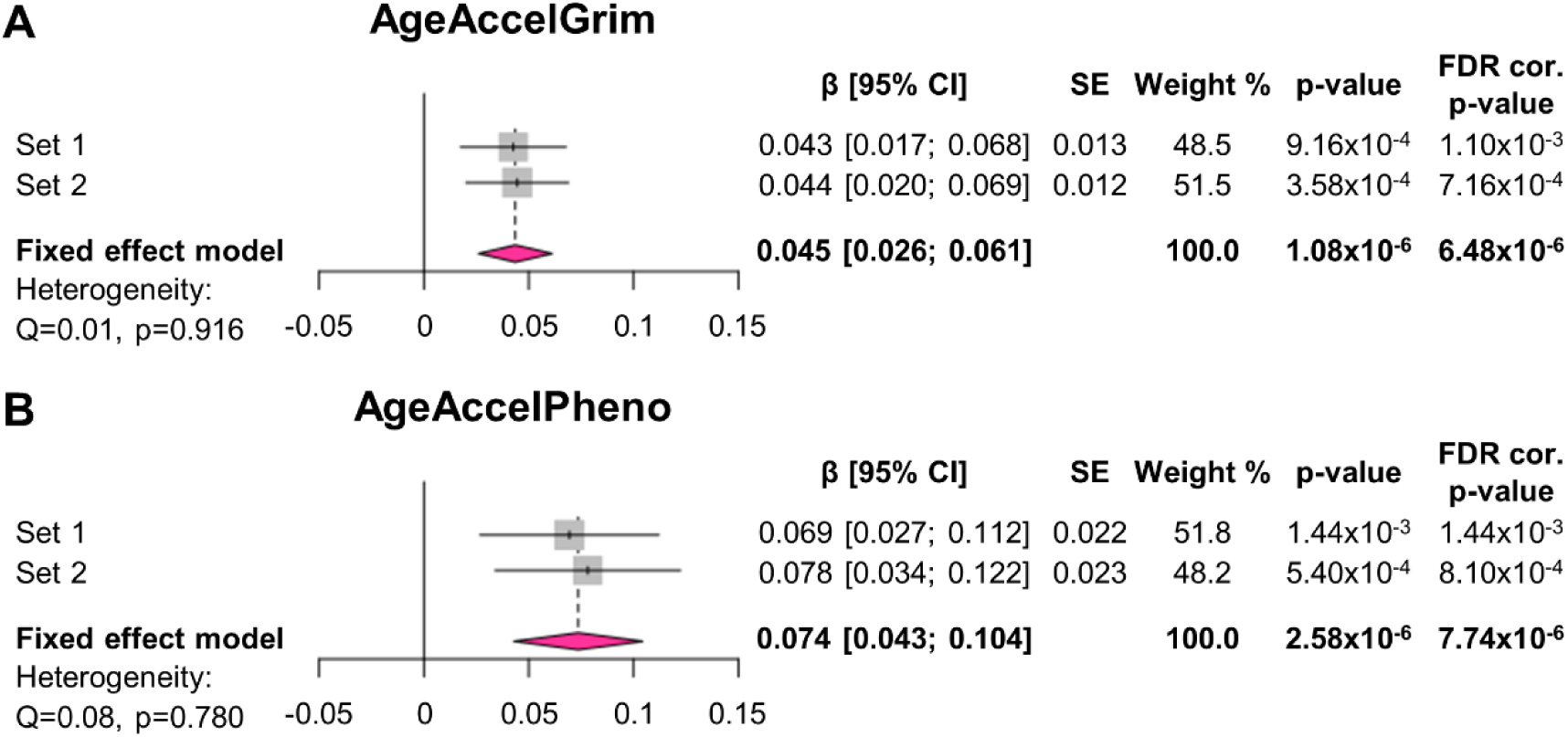
Alcohol consumption is associated with advanced GrimAge and PhenoAge in non-smokers. Effects of alcohol consumption (units/week) on (A) AgeAccelGrim and (B) AgeAccelPheno in non-smoking participants. Values on forest plot indicate standardised **β** with 95% confidence intervals. Models are adjusted for sex and BMI in sets 1 and 2, and relatedness in set 1 by fitting pedigree information as a random effect in general linear mixed models using advanced restricted maximum likelihood (ASReml) method. Fixed effect inverse variance weighted meta-analysis was applied using R package *meta* to combine the standardised coefficient estimates in set 1 and set 2. FDR correction was applied across all smoking sensitivity models in sets 1 and 2, and all meta-analysis models (6 models in total). Sample size: n=2207 in set 1, n=1998 in set 2 (n=4205 included in meta-analyses). EEAA = extrinsic epigenetic age acceleration, IEAA = intrinsic epigenetic age acceleration, SE = standard error, CI = confidence interval, FDR = false-discovery rate.

### Testing for the causal influence of alcohol use on accelerated brain age

Having demonstrated a consistent phenotypic association between alcohol use and accelerated brain age, we used two-sample MR to test a causal relationship between AUD/alcohol consumption (AUDIT-C) and brain age (Figure 4). For AUD, the IVW model was significant (β=0.272 [0.007; 0.537], p=0.044; mF=13.909 (Table S7)) (Figure 4B) but not for AUDIT-C (Figure 4A), suggesting that AUD, but not alcohol consumption levels, has a possible causal effect on accelerated/advanced brain age. However, sensitivity analyses showed that most of the SNPs included in the MR of AUD and brain age were either weak instruments as indicated by F-statistics (Table S7), or violated the assumption of directionality as measured by Steiger filtering (Table S13). Additionally, Radial MR revealed rs570436 (Q=13.409, p=0.011) as an outlier in the MR analysis of AUD and brain age, and therefore the analysis was repeated with this SNP excluded and SIMEX correction applied for the MR Egger analysis, as I^2^ was <0.9 (Table S6). After removing the outlying SNP, the IVW model was no longer significant (β=0.211 [-0.003; 0.425], p=0.054), suggesting that the outlier was driving the significant result for AUD (Figure S3), although the biological function of this variant is not known so we cannot be sure whether it acts through alcohol consumption. Together these results provide weak evidence for a causal effect of genetically instrumented AUD on brain age, and no evidence of a causal effect of alcohol use on brain age as measured by AUDIT-C.

**Figure 4.**
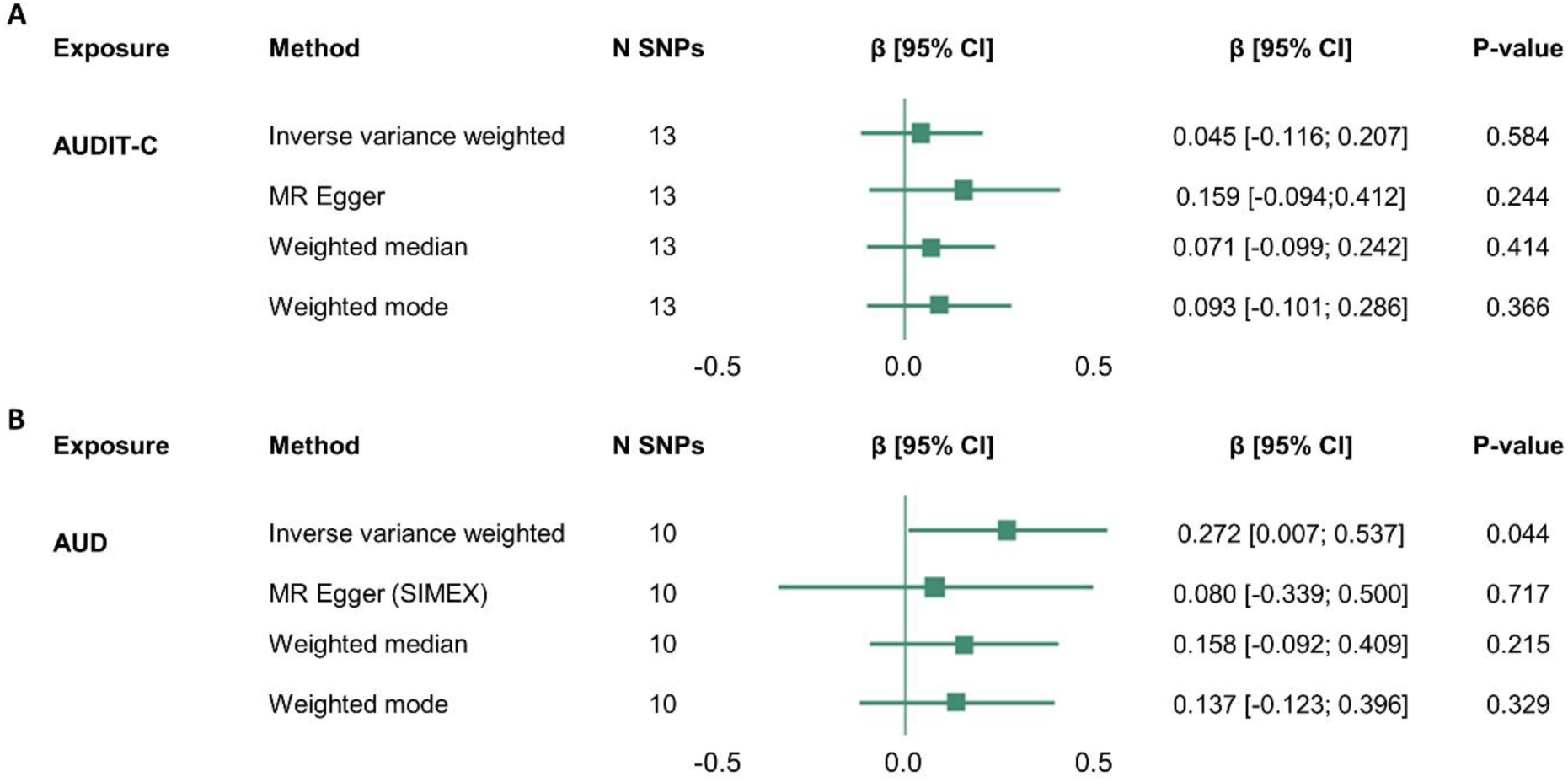
Two sample Mendelian randomisation analysis provides weak evidence for a causal effect of AUD on brain age acceleration. (A) Two sample Mendelian randomisation of AUDIT-C on Brain Age; (B) Two sample Mendelian randomisation of AUD on Brain Age, with SIMEX correction applied to MR-Egger as I^2^ was <0.9. Weighted SIMEX is reported. Data on the genetic association with AUDIT-C and AUD was extracted from Kranzler et al (33). Summary statistics for these SNPs were extracted from a novel GWAS of Brain Age (see Supplementary methods). N SNP=number of SNPs included in the MR analysis, CI = confidence interval, AUD = Alcohol use disorder, SIMEX = simulation extrapolation.

### Testing for the causal influence of alcohol consumption on epigenetic age acceleration

As we observed a significant association between alcohol consumption and advanced GrimAge and PhenoAge, we used two-sample MR methods to test whether these effects might be causal. There was no evidence to suggest a causal effect of alcohol consumption (AUDIT-C) on the two EAA measures (Figure 5). The mean F-statistics suggest that the SNPs included in the analyses are strong genetic instruments (mF=79.058; Table S7). We found little evidence of heterogeneity for the association between AUDIT-C and AgeAccelGrim as assessed by Cochran’s Q statistic while heterogeneity was present for the association between AUDIT-C and AgeAccelPheno (Table S8). However, the MR-Egger intercept suggested that this was not due to directional pleiotropy (Table S9). Steiger filtering also revealed that some SNPs included in the MR analyses explain significantly more variance in the outcome than in the exposure (Table S11-12), violating the assumption of directionality. Together these results show that in this study we find no clear evidence that the association between alcohol consumption and accelerated epigenetic age is causal.

**Figure 5.**
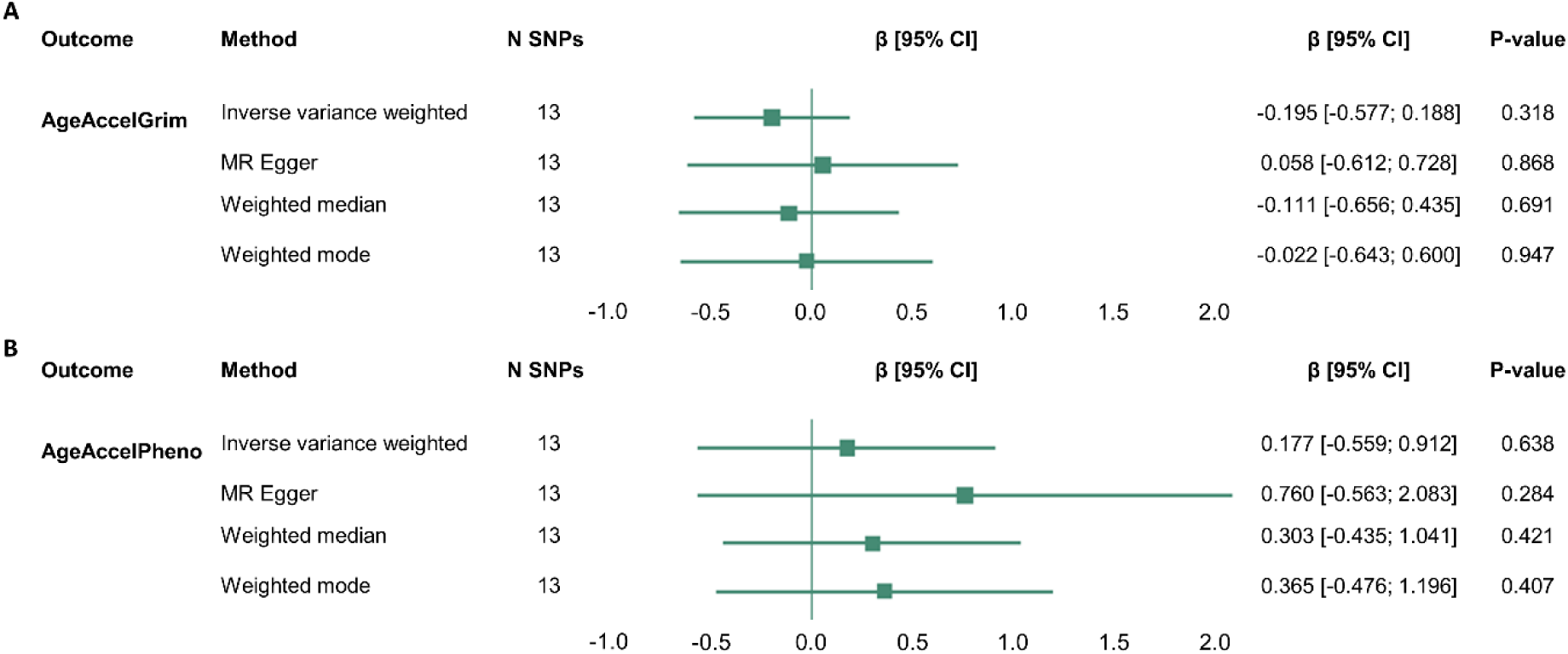
Two sample Mendelian randomisation analysis shows no evidence for causal effect of alcohol consumption on measures of epigenetic age acceleration: (A) AgeAccelGrim and (B) AgeAccelPheno. Data on the genetic association with alcohol use (AUDIT-C) was extracted from Kranzler et al (33). Summary statistics for these SNPs were extracted from GWASs for AgeAccelGrim and AgeAccelPheno conducted by McCartney et al (31). rs185177474 was not available in AgeAccelGrim and AgeAccelPheno summary statistics, thus rs151242810 was used as a proxy (R^2^=0.976; see Supplementary methods).N SNP=number of SNPs included in the MR analysis, CI = confidence interval.

## Discussion

This study represents one of the largest systematic investigations of alcohol use and biological ageing to date, and is the first study investigating possible causal relationships between alcohol use and accelerated brain and epigenetic ageing using two-sample MR. We report consistent positive associations between four measures of alcohol use and accelerated brain age as well as alcohol consumption and two measures of DNAm age acceleration (AgeAccelPheno and AgeAccelGrim). MR analyses revealed limited evidence for the causal effect of AUD on accelerated brain age, while there was no evidence to suggest a causal link between levels of alcohol consumption and brain or epigenetic ageing.

In the present study we demonstrate a positive association between four measures of alcohol use and a metric of accelerated brain ageing, derived from structural MRI. We show associations with problematic alcohol use (AUDIT-P), alcohol consumption (AUDIT-C) and total AUDIT scores, with the strongest phenotypic association found for alcohol consumption in units/week. These results are consistent with previous investigations showing that brain changes associated with ageing are more pronounced in individuals with higher levels of alcohol use (2), and with previous reports of advanced brain age relative to peers in individuals who consume alcohol more frequently (22,27). The present study provides evidence that brain changes in response to excessive alcohol use resemble an early ageing process.

We further demonstrate a positive association between alcohol consumption and accelerated DNAm PhenoAge and GrimAge, thus expanding on the results of Fiorito et al (25) who showed positive association between alcohol consumption levels and accelerated PhenoAge. However, we do not replicate previous findings showing associations between self-reported alcohol consumption levels and accelerated DNAm age derived from Hannum and Horvath clocks (17,25) which could be attributed to differences in study populations (17), differences in the use of alcohol use variables in the models (25), and the use of different DNAm arrays (17,25). Furthermore, the first generation clocks (Horvath and Hannum) were designed for the Illumina 27k and 450k arrays, whereas PhenoAge and GrimAge were developed using overlapping CpG sites on the Illumina 450K and EPIC arrays, and there are mixed findings of whether the Hannum and Horvath age estimations are accurate when using the EPIC array (59,60). Importantly, the novel PhenoAge and GrimAge estimators are shown to be stronger predictors of mortality and lifestyle factors, including alcohol use (25,30,44) which could be explained by the inclusion of CpG sites associated with biomarkers of physiological dysregulation and disease, while Hannum and Horvath clocks were designed to predict chronological age.

A major strength of the current study is the use of data from both UKB and GS:SFHS, which enabled association studies to be conducted in much larger samples compared with previous reports (n=20,258 for brain ageing and n=8,051 for the DNAm ageing). An additional strength is the range of measurements enabling a systematic assessment of the association of four different self-reported alcohol use measures with brain age acceleration as well as the association between alcohol consumption and four different measures of EAA.

We report limited evidence suggesting a causal link between the diagnosis of AUD and accelerated brain ageing, but no clear evidence for higher alcohol consumption causing accelerated brain or epigenetic ageing. We replicate and expand on recent findings showing significant genetic correlations between alcohol-related phenotypes and epigenetic ageing, but no causal relationship between alcohol use frequency and EAA measures as assessed by MR (31). This suggests that the phenotypic association may arise from confounding factors (e.g. other harmful lifestyle factors) that have directional effects on both alcohol use and biological ageing. High levels of psychiatric comorbidity with AUD (61–63) represent another possible confounder. For example, schizophrenia (64), major depressive disorder (65–67) and post-traumatic stress disorder (68) are associated with accelerated brain and/or epigenetic ageing. Furthermore, sensitivity analyses revealed that the genetic instruments used here violate key assumptions of MR analysis including vertical pleiotropy and directionality, suggesting potential reverse causation. Finally, the Million Veteran Programme sample used for the GWAS of AUD and alcohol consumption comprises predominantly male armed forces veterans (33).Thus it may not be representative of the whole population, as there is evidence that alcohol use and AUD has higher prevalence in males and in veterans (1,69), and genetic mechanisms might differ between sexes. Additionally, there are differences in the phenotypes used in the current study and the ones used in the GWAS for AUD and alcohol consumption by Kranzler et al (33) (e.g., clinically diagnosed AUD vs AUDIT-P). Future longitudinal studies combined with more experimental approaches could help elucidate the mechanisms linking alcohol use with biological ageing, and their interactions.

Several other limitations need to be addressed when interpreting these findings. First, this study relied on self-reported measures of alcohol use that might be inaccurate due to response-biases. Whereas there are traditionally few alternatives, the validation of approaches such as estimation of alcohol drinking via the generation of a composite score for alcohol use from DNAm data (26,70), or other biological data (e.g. liver enzyme levels as investigated in association with EAA previously (24)) could help to overcome the need to rely on self-reported measures. Second, although we investigated the associations between problematic alcohol use (AUDIT-P) and brain ageing, we were unable to conduct a similar evaluation in association with DNAm ageing measures. Additionally, different drinking patterns may be important. It was recently suggested that the relationship between educational attainment and adverse health outcomes is mediated by specific patterns of alcohol use, such as binge drinking, rather than total alcohol consumption (71). Thus, the focus on overall alcohol consumption in the present study might preclude the detection of a causal relationship. Finally, we investigated the associations between alcohol use and brain ageing or blood DNAm ageing in two separate large cohorts. It would be of interest in future studies to investigate the associations between alcohol use and the two types of biological ageing measures in the same individuals and evaluate the relationship between brain and DNAm ageing.

To conclude, in one of the largest studies on the relationship between alcohol use and biological ageing to date, and a first investigation of the causal relationship between the two using MR, we report a consistent association between higher levels of alcohol consumption and accelerated biological ageing. The present study found some evidence that the diagnosis of AUD might be causally linked to accelerated brain ageing, but no clear evidence for a causal link between alcohol consumption levels and biological ageing. The positive phenotypic associations between alcohol consumption and brain and epigenetic ageing add to the body of literature suggesting that alcohol use may exert a detrimental effect on the body’s vital organs in a way that resembles early ageing. This suggests that accelerated biological ageing may mediate the association of alcohol use with age-related disease and increased mortality.

## Data Availability

According to the terms of consent for GS:SFHS, access to data must be reviewed by the GS Access Committee.
Details concerning access to UK Biobank data can be found in the link provided.
https://www.ukbiobank.ac.uk/principles-of-access/
Summary statistics from GWAS of AgeAccelPheno and AgeAccelGrim available here: https://datashare.is.ed.ac.uk/handle/10283/3645
Summary statistics from GWAS of brain age acceleration available here: https://datashare.is.ed.ac.uk/handle/10283/3797

## Acknowledgments

This work was supported by a Wellcome Trust Strategic Award “Stratifying Resilience and Depression Longitudinally” (STRADL) [104036/Z/14/Z]. Generation Scotland received core support from the Chief Scientist Office of the Scottish Government Health Directorates [CZD/16/6] and the Scottish Funding Council [HR03006]. Genotyping of the GS:SFHS samples was funded by the Medical Research Council UK and the Wellcome Trust (Wellcome Trust Strategic Award (STRADL; Reference as above). DNA methylation profiling of the GS:SFHS samples was funded by the Wellcome Trust Strategic Award [10436/Z/14/Z] with additional funding from a 2018 NARSAD Young Investigator Grant from the Brain & Behavior Research Foundation [27404]. The UK Biobank core activities and imaging study was funded primarily by the Wellcome Trust and the Medical Research Council (MRC), as well as the Department of Health and the National Institute for Health Research (NIHR), the Scottish and Welsh Governments, the British Heart Foundation and Cancer Research UK. SMKB and KV are funded by the Wellcome Trust Translational Neuroscience PhD Programme at the University of Edinburgh [108890/Z/15/Z]. JHC is supported by a UKRI Innovation Fellowship (MR/R024790/2). SRC received support from the UK MRC [MR/R024065/1] and US National Institutes of Health [R01AG054628]. DLM and REM were supported by Alzheimer’s Research UK Major Project grant (ARUK-PG2017B-10). REW works in a unit that receives funding from the University of Bristol and the UK Medical Research Council (MC_UU_00011/1 and MC_UU_00011/3).

