## Supplemental methods and figures for "Associations between alcohol use and accelerated biological ageing"

### Supplementary methods

#### DNA methylation profiling

Whole blood genomic DNA samples were treated with sodium bisulphite using the EZ-96 DNA Methylation Kit (Zymo Research, Irvine, California), following the manufacturer's instructions. Genome-wide DNA methylation (DNAm) was measured using the Infinium MethylationEPIC BeadChip (Illumina Inc., San Diego, California) in accordance with the manufacturer's protocol. DNAm was profiled in two sets (set 1 n=5,191 and set 2 n=4,583) at separate time points and quality control and normalisation was carried out separately. Arrays were scanned using a HiScan scanner (Illumina Inc., San Diego, California), with initial inspection of array quality carried out in GenomeStudio v2011.1. Additional quality control measures were implemented using the R packages 'shinyMethyl' (1), meffil (2) and 'wateRmelon' (3) as described in detail previously (4,5). Briefly, outlier sites and participants, together with participants for whom there was a mismatch between their predicted sex (based on DNAm data) and their recorded sex, were excluded from both sets; samples were then normalised (separately) using the dasen method from the wateRmelon R package. The final DNAm dataset comprised data for 5087 individuals in set 1 and 4450 individuals in set 2.

#### Epigenetic estimates of age

Epigenetic age and age acceleration measures were calculated using the online age calculator (https://dnamage.genetics.ucla.edu/) developed by Horvath (6). Normalised DNAm beta-values were submitted to the calculator, using the 'Advanced Analysis for Blood Data' option.

The Horvath epigenetic age was calculated based on methylation levels at 353 CpG sites following the approach developed by Horvath (6). Hannum epigenetic age was calculated based on DNAm levels at the 71 CpGs identified by Hannum (7). From these DNAm age estimates we derived two measures for epigenetic age acceleration that is either independent of blood cell counts or enhanced by changes in blood cell composition, as described previously (8–10). In brief, intrinsic epigenetic age acceleration (IEAA) is defined as the residual term of a multivariate model regression estimated Horvath methylation age on chronological age, adjusting for blood cell counts estimated from the methylation data (naive CD8+ T-cells, exhausted CD8+ T-cells, plasmablasts, CD4+ T-cells, natural killer cells, monocytes, and granulocytes), and is therefore by definition independent of blood immune cell counts. Extrinsic epigenetic age acceleration (EEAA), on the other hand, tracks age-related changes in blood cell composition as well as cell-intrinsic epigenetic changes. It is estimated by first calculating Hannum's DNA methylation age; and second, increasing the contribution of three cell types whose abundance is known to change with age (naive cytotoxic T-cells, exhausted cytotoxic T-cells, and plasmablasts) by forming a weighted average of Hannum's DNA methylation age estimate with these three cell type estimates using the Klemera-Doubal approach (11). EEAA is defined as the residual term of univariate model regressing the weighted estimated Hannum's epigenetic age in chronological age.

The DNAm GrimAge was calculated using a method developed by Lu et al (12) and is based on a linear combination of age, sex, DNAm-based surrogates for smoking, and seven proteins (adrenomedulin, beta-2-microglobulin, cystatin C, growth differentiation factor 15, leptin, plasminogen activation inhibitor 1, and tissue inhibitor metalloproteinaise). Adjusting DNAm GrimAge for chronological age generated the measure of epigenetic grim age acceleration, AgeAccelGrim (12).

The DNAm PhenoAge was calculated using a method developed by Levine et al (13) and is based on a weighted average of ten clinical characteristics (chronological age, albumin, creatinine, glucose and C-reactive protein levels, lymphocyte percentage, mean cell volume, red blood cell distribution width, alkaline phosphatase and white blood cell count) regressed on DNAm levels. This approach generates the selection of 513 CpGs and linear combination of the weighted CpGs yields the DNAm PhenoAge (13). Adjusting DNAm PhenoAge for chronological age generated the measure of phenotypic epigenetic age acceleration, AgeAccelPheno.

**Two-sample Mendelian randomisation**

*Exposure GWAS: Alcohol consumption (AUDIT-C) and Alcohol Use Disorder (AUD).* Data on the genetic association with alcohol use was extracted from Kranzler et al (14). This study was carried out in the Million Veterans Program population, which is based in the USA and therefore highly unlikely to overlap with UKB or GS:SFHS populations made up of British and Scottish individuals, respectively. Kranzler et al (14) reported a genome-wide significant association between 10 single nucleotide polymorphisms (SNPs) and AUD, as well as 13 SNPs associated with alcohol consumption as measured by AUDIT-C. These SNPs were identified as independent by linkage disequilibrium (LD) clumping using a 500-kb genomic window and $r2 < 0.1$ (14). Here, we extracted the summary statistics as reported in the European Ancestry population (see Supplementary Table 1 and 2 in Kranzler et al (14)).

*Outcome GWASs*:

*Brain Age:* GWAS for brain age acceleration was performed in a subset of unrelated White British individuals in the UKB imaging cohort. 16,133 individuals were included, using a relatedness cutoff of 0.05. To perform the GWAS, brain age was entered as the outcome variable and age, sex, genotyping array, and the 20 first principle components derived from genotype data as covariates. From the results of this analysis we extracted the summary statistics (beta and standard error of the effect allele of each SNP) for the 10 and 13 SNPs identified as genome-wide significant by Kranzler et al (14) for AUD and AUDIT-C, respectively. The full GWAS summary statistics are available here: https://datashare.is.ed.ac.uk/handle/10283/3797.

*Epigenetic Age:* Data on the genetic association with AgeAccelGrim and AgeAccelPheno was extracted from McCartney et al (15). These GWASs were conducted in 34,962 European ancestry individuals and a fixed-effect meta-analysis was performed to combine the summary statistics (15). Supplementary tables were inspected to ensure that the Million Veteran Program cohort was not included in these meta-GWASs. From the GWAS results (available here:

https://datashare.is.ed.ac.uk/handle/10283/3645) we extracted the corresponding summary statistics for the 13 SNPs identified as GW significant for AUDIT-C by Kranzler et al (14).

For SNPs unavailable in the outcome GWAS summary statistics, proxy SNPs were searched for (https://ldlink.nci.nih.gov/?tab=ldproxy) with a minimum LD $R^2$=0.9. In AgeAccelGrim and AgeAccelPheno GWASs, rs185177474 was not available, thus rs151242810 was used as a proxy ($R^2$=0.976).

See Tables S4 and S5 for the full MR input testing causal effects of AUDIT-C and AUD, respectively, on biological ageing.

### Supplementary Figures

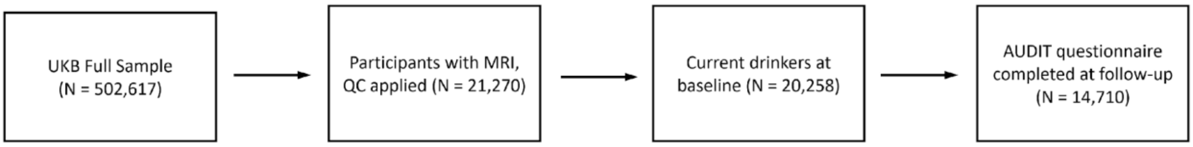


**Figure S1.** Flowchart showing sample selection in UKB for the present study. At the time of analysis, the number of participants in UKB totalled N=502,617 and of these, the latest neuroimaging release consisted of N=21,270 after QC application (see main text for details of QC protocol). Of these, the present study included N=20,258 individuals who reported being current drinkers. A subset of UKB participants completed an online follow-up including the AUDIT questionnaire. Of the present sample of current drinkers with available MRI data, N=14,710 had completed the AUDIT questionnaire.

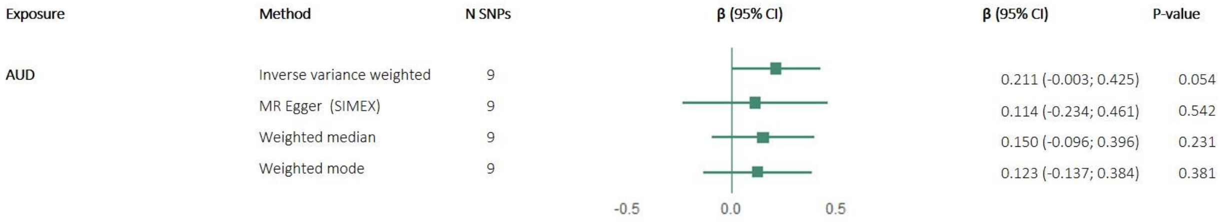


**Figure S2.** Two sample Mendelian randomisation of AUD on brain age. Results of Mendelian randomisation analysis with the outlying SNP rs570436 (Q=13.409, p=0.011, Radial MR) removed from the main analysis (Figure 4). SIMEX correction was applied to MR-Egger as $I^2$ was <0.9. Weighted SIMEX-corrected estimate is reported. Data on the genetic association with AUD was extracted from Kranzler et al (14). Summary statistics for these SNPs were extracted from a novel GWAS of Brain Age (see Supplementary methods). N SNP=number of SNPs included in the MR analysis. No further sensitivity analysis was applied here.
